# Orthostatic hypotension in Parkinson’s disease impacts the association between white matter lesion volume and motor symptom burden

**DOI:** 10.64898/2026.01.25.26344797

**Authors:** John D’Amico, Miriam Sklerov, Eran Dayan

## Abstract

**Objective:** To evaluate whether the presence of orthostatic hypotension (OH) impacts the association between white matter hyperintensity (WMH) volume and motor symptom burden in persons with Parkinson’s disease (PWP).

**Methods:** Motor symptom burden in PWP was quantified using the Movement Disorders Society Unified Parkinson’s Disease Rating Scale (MDS-UPDRS) Part III. Total WMH volume was segmented based on high-resolution T1-weighted (T1W) and T2 Fluid Attenuated Inversion Recovery (FLAIR) images. Determination of whether individual participants qualified as having OH was based on orthostatic vital signs. All data were obtained from the Parkinson’s Progression Markers initiative (PPMI) dataset.

**Results:** A total of 218 PWP (mean age, 64.84 ± 9.51) and 50 control participants (mean age, 65.52 ± 11.04) were included in the analyses. WMH volume did not differ significantly between the two groups. 15.1% of the participants in the PD group and only 4% of the control group qualified as having OH. Analysis of covariance determined that in PWP, the association between WMH volume and motor symptom burden was significantly different in participants with OH in comparison to those without OH (steeper in the former group). Follow-up analyses suggested that the effects were strongest for bradykinesia symptoms. Adjusting for disease and symptom duration did not alter results.

**Conclusions:** The presence of OH in PWP impacts the links between white matter lesion volume and motor symptom burden. These findings may provide a potential mechanism underlying the poorer disease prognosis among PWP with OH.

## Introduction

Parkinson’s disease (PD) is a complex movement disorder that primarily results in the cardinal motor symptoms of bradykinesia, tremor, rigidity and postural instability^1^. It is by now widely acknowledged that PD is also associated with a collection of non-motor symptoms^2^, such as cognitive decline^3^, sleep disturbances^4,5^, sensory dysfunction^6^, and autonomic impairments^7,8^. Associations between motor and non-motor symptoms in persons with PD (PWP), and their mechanistic origin remain a topic of continued interest.

The brain mechanisms that underlie the motor and non-motor symptoms of PD are complex and multifaceted^9^. Among multiple molecular, cellular and circuit mechanisms, white matter lesions – which appear as hyperintense signals on T2 magnetic resonance imaging (MRI) – have also been studied^10^, primarily in relation to cognitive decline in PD^11,12,13^. In contrast, less is known about the associations of these so-called white matter hyperintensities (WMH) with motor symptoms of PD, with inconsistent findings reported in the literature^14,15,16^. The discordance between the findings of prior studies suggests that there are certain factors that may influence the relationship between WMH volume and motor symptom burden in PWP, however, these factors remain mostly unknown to date.

A plausible factor that could impact the association between WMH volume and motor symptom burden in PD is autonomic dysfunction, particularly orthostatic hypotension (OH). OH has been shown to be linked with poor health outcomes^17^ and, along with subjective cardiovascular autonomic symptoms, to have an adverse effect on activities of daily living^18^ and quality of life^17,19^. Importantly, associations between OH and WMH volume were reported in both older adults^20^ and PWP^21^. Whether OH contributes to the relationship between WMH volume and motor symptom burden in PD has yet to be determined.

Here, using the data from the Parkinson’s Progression Markers Initiative (PPMI), we investigated whether the presence of OH in PWP impacts the association between WMH volume and impairment of motor function (Fig. 1). The volume of WMH was quantified using T1W and T2 FLAIR images, while motor symptom burden was assessed with the Movement Disorder Society Unified Parkinson’s Disease Rating Scale (MDS-UPDRS) Part III. The presence of OH was categorized using available clinical vital signs and previously established cutoffs^22,23^. Analyses then determined the extent to which OH affects the magnitude of associations between WMH volume and motor symptom burden, whether these effects covaried with measures of disease duration and disease severity, and the specific motor symptom domains most strongly implicated in these associations.

**Figure 1:**
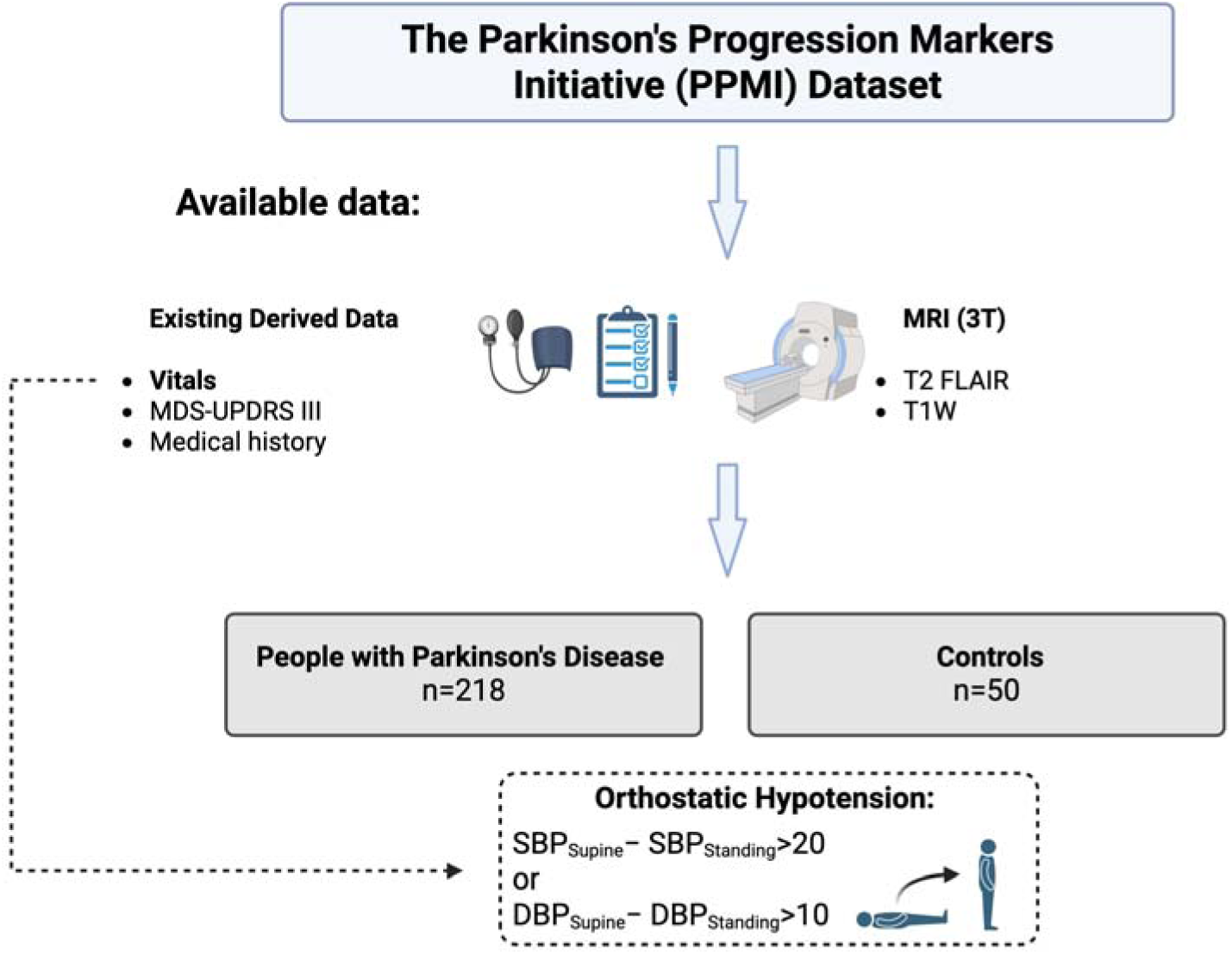
Study design. All data were obtained from the PPMI dataset. T1W and T2 FLAIR magnetic resonance images from 268 individuals were used, along with their corresponding medical history, vital signs, and Movement Disorder Society-Unified Parkinson’s Disease Rating Scale Part III (MDS-UPDRS III) scores. We determined if each participant had OH based on whether their systolic or diastolic blood pressure changed by 20 or 10 points, respectively, when moving from supinated to standing positions. We hypothesized that the presence of OH in participants will significantly impact the association between total WMH volume and motor symptom burden in PWP. DBP: diastolic blood pressure, SBP: systolic blood pressure.

## Methods

### Participants

All data were obtained from the PPMI dataset, a comprehensive, observational multi-center study of early PD. In the current study, we analyzed data from the PPMI’s main PD and healthy controls cohorts. Full inclusion and exclusion criteria for these and other cohorts in the PPMI can be accessed online (www.ppmi-info.org). In short, to be included in the PD cohort, participants had to be age 30 or above, have a diagnosis of PD ≤ 2 years from the time of screening, Hoehn and Yahr stage I or stage II, supportive positron emission tomography or Dopamine Transporter (DaT) scan findings, and at least two of the following symptoms: resting tremor, bradykinesia, or rigidity. Participants were expected to not have been on a PD medication regiment within 6 months of their baseline (BL) visit. Control participants were excluded if they had first degree relatives with PD, or if they had a significant neurological or psychiatric disorder. All procedures were approved by the Institutional Review Boards of the participating sites, and all participants provided written informed consent prior to enrolling in the study.

Data included in all analyses reported here were limited to participants’ earliest visits containing the variables of interest (see below). We reasoned that limiting our data to the initial visit for participants would minimize the impact of confounding factors (i.e. comorbidities, clinical complications, and other related factors). To select participants for analyses, we first identified both PWP and controls who had T1W, T2 FLAIR and MDS-UPDRS Part III exam scores available at the same visit. Therefore, the final sample consisted of 218 PWP (mean age: 64.84 years ± 9.51; all were not receiving PD medications at the time of testing) and 50 controls (mean age: 65.52 ± 11.04).

### Imaging Data

All imaging data were acquired with Siemens 3T scanners. The current study utilized T1W and T2 FLAIR images. High-resolution T1W scans were acquired with a magnetization prepared rapid acquisition gradient echo (MPRAGE) sequence (TR = 2300ms, TE = 2.98, 1.0 mm slice thickness, 192 slices, 1.0 x 1.0 mm voxel size, 256 x 256 matrix, FOV = 256 mm). 3D T2 FLAIR sagittal images were also acquired as well (1.0-1.2 mm, 192 slices, 1.0 x 1.0 mm voxel size, FOV = 256 mm). Additional relevant technical information can be obtained on the PPMI website (https://www.ppmi-info.org/study-design/research-documents-and-sops).

### Scoring of Motor Function

Participants’ motor symptom burden was quantified using MDS-UPDRS Part III scores collected at the PPMI BL visits. Those included in the current study were not on medications to treat motor symptoms of PD. Part III of the MDS-UPDRS is a motor examination which includes 18 items, focusing on different aspects of motor symptom burden for PWP; higher scores indicate worsened motor function in participants. Score ranges for part III are 0 to 132. The reported Hoehn and Yahr (H&Y) stages were additionally extracted for each participant. Calculation of motor domain sub-scores were completed as well based on the MDS–UPDRS Part III items. Tremor (UPDRS items 20 and 21), rigidity (UPDRS item 22), bradykinesia (UPDRS items 23-26), and axial symptoms (UPDRS items 27-30) sub-scores were thus summed for each participant^14^.

### Definition of Orthostatic Hypotension

Determination of OH was based on orthostatic vital signs data included in the PPMI. Blood pressure (BP) and heart rate were initially measured while the participants were in the supine position and then measured again 3 minutes after being moved to the standing position. We defined OH as a change of ≥ 20 mmHg in the participants’ systolic BP reading, or a ≥ 10 mmHg change in their diastolic BP reading, within 3 minutes of standing^22^. Of note, these criteria are aligned with the American Academy of Neurology Consensus Statement on OH^23^.

### Measures of Disease Duration

Reported time from diagnosis (disease duration) and time from the onset of symptoms (symptom duration) were extracted for all PWP. Since our focus here was on early PD, both variables had restricted ranges. Thus, as an additional measure of disease severity, we utilized DaT scan data, available on the PPMI database as derived variables. For each participant, we calculated mean DaT scores by averaging DaT availability in the caudate and putamen: the main nuclei to the basal ganglia^24^.

### Medical Comorbidities

Medical comorbidities were categorized into 17 categories in the PPMI dataset: such as cardiovascular, musculoskeletal, neurologic and gastrointestinal. We considered their impact in follow-up analyses.

### Data analysis

Calculation of WMH volume for all participants was completed using the lesion growth algorithm^25^ in the Lesion Segmentation Toolbox (https://www.applied-statistics.de/lst.html), with default settings, running on MATLAB R2024b (MathWorks, Natick, MA). Total volume of WMH was then log-transformed, due to a skewed distribution across participants^26^. The estimated total intracranial volume (eTIV) of participants was additionally obtained from MPRAGE data using FreeSurfer version 8.0.0 (https://surfer.nmr.mgh.harvard.edu/). Imaging and non-imaging data were all matched by study visit.

Group differences in continuous variables were evaluated with independent sample t-tests, while differences in categorical variables were evaluated with χ^2^ tests. Our main analysis tested whether the association between WMH volume and MDS-UPDRS Part III scores was impacted by the presence of OH. This was examined using an analysis of covariance (ANCOVA) model, testing for the effect of the interaction between OH and WMH volume on MDS–UPDRS Part III symptom sub-scores. We used a similar approach to test whether OH impacted the association between WMH volume and specific motor symptom domain scores, including bradykinesia, axial, tremors and rigidity. All statistical analyses were performed using JASP version 0.95. Statistical significance was set at a p-value of 0.05.

## Results

### Participant Characteristics

Our objective in this study was to examine whether the association between WMH volume and motor symptom burden in PWP is impacted by the presence of OH. We analyzed data from a sample of 218 PWP and 50 controls. The two groups did not significantly differ in age, sex, supine or standing diastolic and systolic BP, WMH volume, or eTIV (Table 1).

**Table 1:** Descriptive and inferential statistics of demographics in included PD participants.

|  | Group | N | Mean | SD | p |
| --- | --- | --- | --- | --- | --- |
| Age | PD | 218 | 64.839 | 9.511 | 0.659 |
|  | PD: OH– | 185 | 64.054 | 9.522 |  |
|  | PD: OH+ | 33 | 69.242 | 8.280 |  |
|  | Control | 50 |  |  |  |
| Sex (M/F) | PD | 218 (128/90) | – | – | 0.168 |
|  | Control | 50 (24/26) | – | – |  |
| Systolic Supine | PD | 216 | 131.556 | 18.534 | 0.479 |
|  | PD: OH– | 183 | 129.366 | 17.262 |  |
|  | PD: OH+ | 33 | 143.697 | 20.824 |  |
|  | Control | 50 |  |  |  |
| Diastolic Supine | PD | 216 | 76.306 | 10.121 | 0.431 |
|  | PD: OH– | 183 | 75.508 | 9.913 |  |
|  | PD: OH+ | 33 | 80.727 | 10.272 |  |
|  | Control | 50 |  |  |  |
| Systolic Stand | PD | 216 | 127.917 | 18.033 | 0.136 |
|  | PD: OH– | 183 | 129.049 | 17.338 |  |
|  | PD: OH+ | 33 | 121.636 | 20.667 |  |
|  | Control | 50 |  |  |  |
| Diastolic Stand | PD | 216 | 78.838 | 11.205 | 0.924 |
|  | PD: OH– | 183 | 79.885 | 10.701 |  |
|  | PD: OH+ | 33 | 73.030 | 12.300 |  |
|  | Control | 50 |  |  |  |
| WMH Volume<br>(log) | PD | 218 | -0.246 | 0.844 | 0.693 |
|  | PD: OH– | 185 | -0.296 | 0.822 |  |
|  | PD: OH+ | 33 | 0.035 | 0.920 |  |
|  | Control | 50 |  |  |  |
| eTIV (mm<sup>3</sup>) | PD | 218 | 1.562E6 | 1.742E5 | 0.178 |
|  | PD: OH– | 185 | 1.561E6 | 1.656E5 |  |
|  | PD: OH+ | 32 | 1.571E6 | 2.201E5 |  |
|  | Control | 50 |  |  |  |

eTIV = estimated total intracranial volume

### Difference in Orthostatic Hypotension Between Groups

Clinical vital signs data were used next to detect OH in all study participants. Changes in BP readings when moving from supine to standing positions were computed for both systolic (Δsystolic_supine_ – Δsystolic_standing_) and diastolic (Δdiastolic_supine_ – Δdiastolic_standing_) BP in the PD (Fig. 2A) and control groups (Fig. 2B). The rates of OH were significantly higher in the PD group (χ^2^_1_ = 4.443, *p* = 0.035). Namely, 15.1% of PWP (n = 33) and only 4.0% (n = 2) of controls qualified as having OH (Fig. 2B).

**Figure 2:**
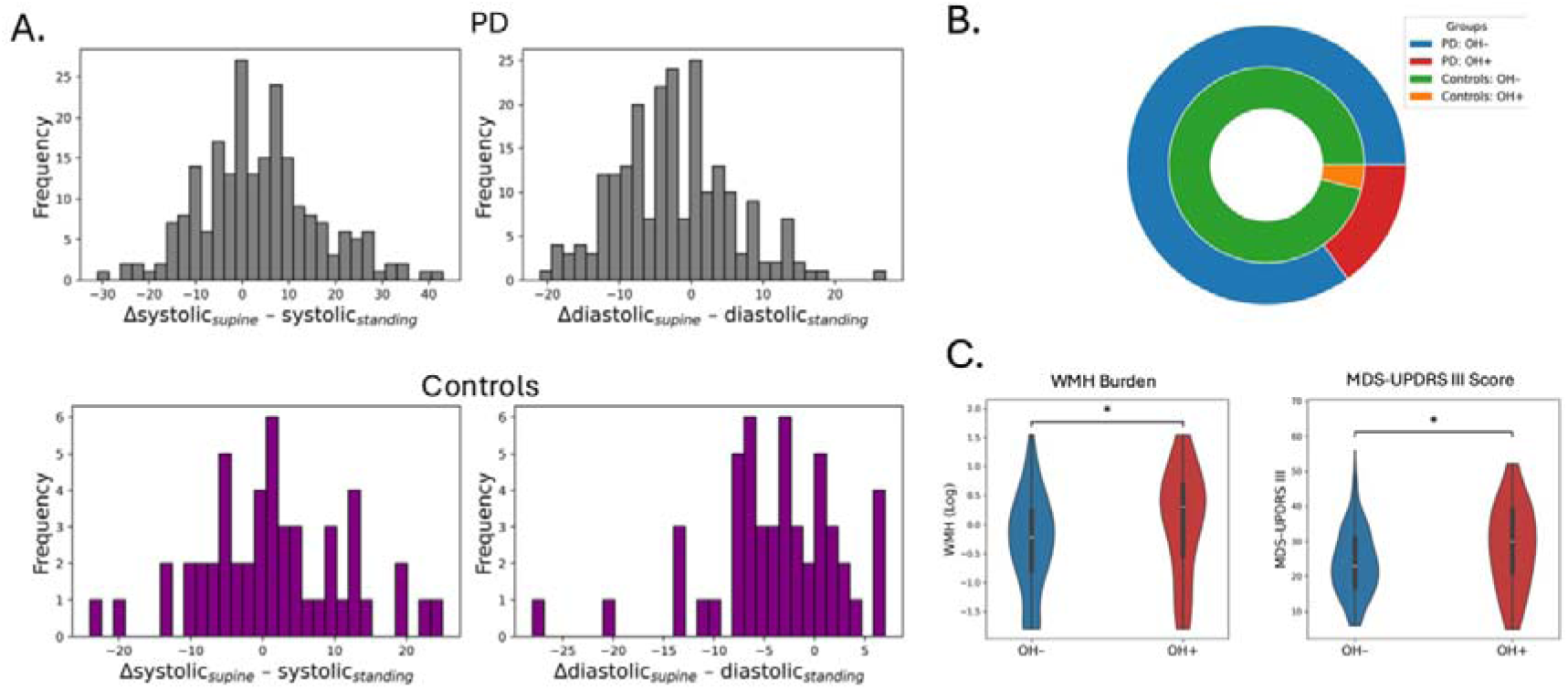
Effect of orthostatic hypotension on white matter hyperintensity volume and motor symptom burden. (A.) Distribution of systolic and diastolic blood pressure drops when moving from supine to standing positions are shown for the PD and control groups. (B.) A total of 33 out of 251 participants in the PD group (15.1%) and 2 participants in the control group (4%) qualified as having OH. (C.) Total WMH volume and motor symptom burden (MDS–UPDRS part III scores) were both higher in PWP with OH (OH+) as compared to those without OH (OH–). * corresponds to *p* < 0.05

### The Impact of OH on the Association Between WMH Volume and Motor Symptom Burden

We then categorized PWP participants into 2 groups based on the presence (OH+) or absence (OH–) of OH. The OH+ group had significantly higher WMH volume (t_216_ = 2.092, *p* = 0.038, Fig 2C), as well as significantly higher MDS–UPDRS Part III total scores (t_215_ = 2.459, *p* = 0.015, Fig. 2C). On the other hand, the OH+ and OH– groups did not differ in 17 categorically coded (1/0) comorbidities (Supplementary Table 1), minimizing potential confounds in subsequent analyses.

Next, we sought to determine the impact of OH on the association between WMH volume and motor symptom burden. In the full sample of PWP, correlations between WMH volume and total MDS–UPDRS Part III scores were significant (*p* = 0.032, Supplementary Table 2), and the strength of these associations differed between OH+ (*p* = 0.027, Supplementary Table 3) and OH– (*p* = 0.414, Supplementary Table 4). We thus next assessed whether the association between WMH volume and motor symptom burden differed in the presence of OH by fitting the data with an ANCOVA model, further adjusting for sex, age, and eTIV. Main effects for neither OH status (F_1,210_ = 3.717, *p* = 0.055) nor WMH volume (F_1,210_ = 2.102, *p =* 0.149) were significant. The OH × WMH volume interaction, however, was significant (F_1,210_ = 4.977, *p* = 0.027, Supplementary Table 5), reflecting a steeper association between WMH volume and MDS–UPDRS Part III total scores in OH+ participants (Fig. 3A).

**Figure 3:**
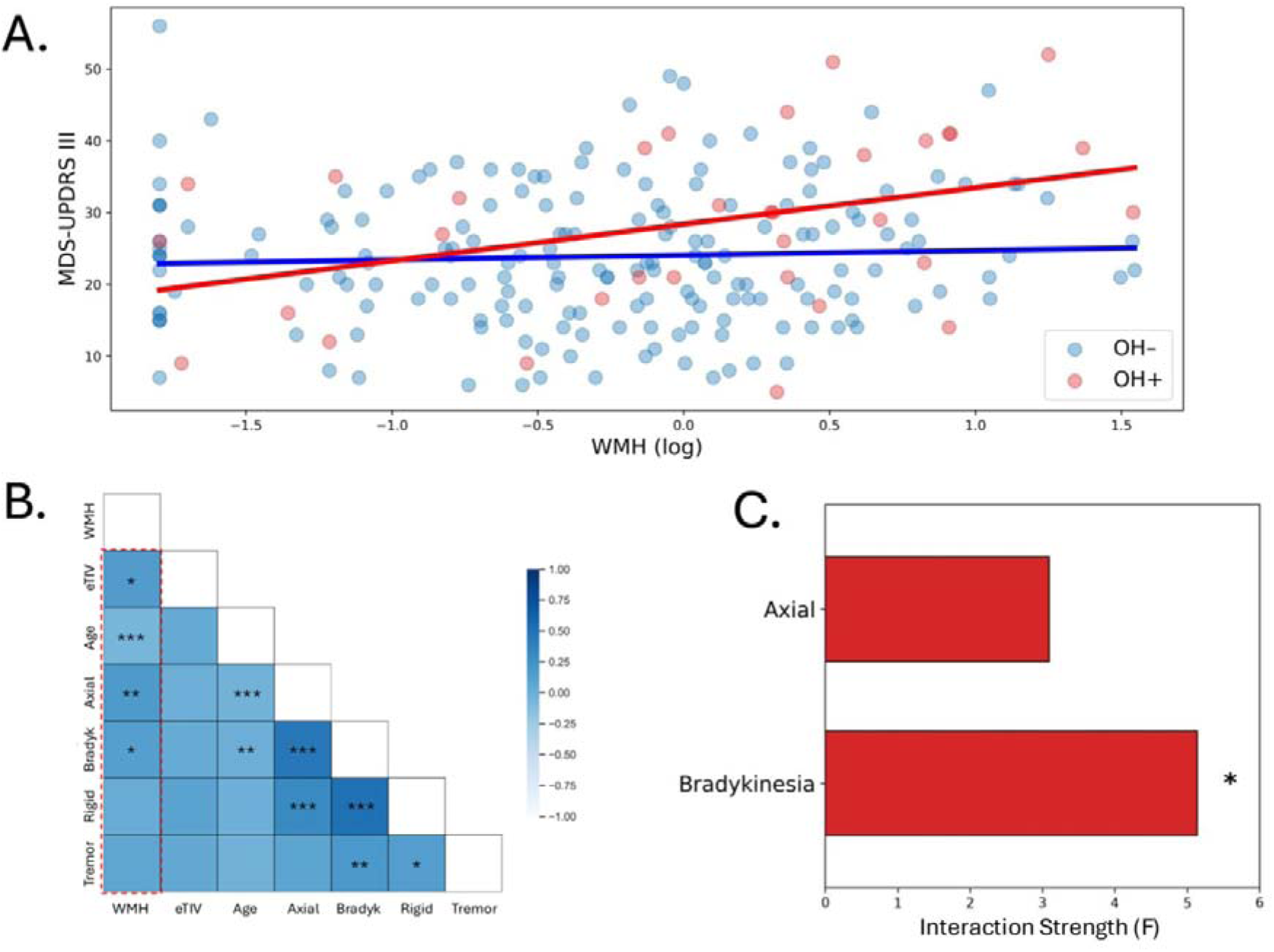
Impact of orthostatic hypotension on associations between white matter hyperintensity volume and motor symptom burden. (A.) An ANCOVA, adjusting for age, sex, and eTIV revealed significant differences in the association between total WMH volume (log-transformed) and motor symptom burden (MDS–UPDRS part III scores) in PWP with OH (OH+) in comparison to those without OH (OH–). (B.) Total WMH volume showed differential correlations with the sub-scores of the MDS–UPDRS part III exam, with a stronger correlation shown for bradykinesia (Bradyk; *p* = 0.050) and axial (*p =* 0.010) sub-scores. (C.) The presence of OH significantly impacted the association between total WMH volume (log-transformed) and bradykinesia sub-scores (*p* = 0.017). A smaller effect was observed for axial sub-scores (*p* = 0.185). * corresponds to p ≤ 0.05

### Effects of Symptom Duration and of Disease Duration and Severity

We next examined if the inclusion of estimated symptom and disease durations of participants affected any of the reported results. Similar OH × WMH volume effects were observed when additionally adjusting for disease (F_1,207_ = 4.787, *p* = 0.030, Supplementary Table 6) and symptom (F_1,207_ = 4.876, *p* = 0.028, Supplementary Table 7) duration. Disease and symptom durations considered here were both narrow due our analysis of BL data and PPMI’s focus on de novo PD. We thus also considered average DaT availability in the striatum (combined putamen and caudate regions of interest), as extracted from DaT scans, as an additional measure of disease severity. Inclusion of this variable as a covariate again resulted in a similar OH × WMH volume interaction (F_1,209_ = 5.971, *p* = 0.015, Supplementary Table 8).

### Analysis of MDS–UPDRS Part III Sub-scores

Our investigation thus far has considered MDS–UPDRS Part III total scores, which may mask the motor domains which drive the reported associations. We thus next sought to examine whether OH impacts the associations between WMH volume and individual motor domain sub-scores. Of the four motor domains considered, WMH volume correlated significantly with axial (*p* = 0.010, corrected for multiple comparisons) sub-scores and showed a marginally significant correlation with bradykinesia (*p* = 0.050, Fig. 3B). We thus repeated the ANCOVA analyses as reported above, focusing on axial and bradykinesia sub-scores as the dependent variables. We found a significant OH × WMH volume interaction for MDS–UPDRS Part III bradykinesia sub-scores (F_1,210_ = 5.141, *p* = 0.024, Supplementary Table 9, Supplementary Fig. 1), but not for axial sub-scores (F_1,210_ = 3.091, *p* = 0.08, Supplementary Table 10, Supplementary Fig. 2).

## Discussion

We report that the effect of WMH volume on the burden of motor symptoms in de novo PD is impacted by the presence of OH. In PWP, OH impacted the strength of the association between WMH volume and MDS–UPDRS Part III scores, with a steeper slope observed in PWP, relative to those without OH. These differential associations were not affected by measures of disease duration, or disease severity. Secondary analyses revealed that the differential impact of OH on links between WMH volume and motor symptom burden were primarily related to bradykinesia symptoms.

We found no group-level differences in WMH volume when comparing controls and PWP, which is in line with previous findings^27,28^; however, the literature is overall inconsistent, with an increased WMH volume in PD reported in several studies^29,30^. It is possible that group-level differences in WMH were not observed in the current study as our focus was on a population of early PD, which may be too early to detect differences in WMH volume compared with controls. Our findings additionally demonstrated that, in comparison to controls, there is a higher proportion of PWP who qualified as having OH, despite early disease. This is consistent with previous research, which consistently demonstrated a higher prevalence of OH among PWP^31,32^.

Previous studies have shown that global^33,34^ or regional^14,35^ WMH volume is associated with worsened parkinsonian motor symptoms, as quantified by MDS–UPDRS Part III score. Our results demonstrate that the strength of these associations is impacted by the presence of OH, with there being a steeper association observed in PWP with OH, compared to those without OH. The mechanisms that underlie these differential associations are not clear at this stage but may reflect the impact of WMH burden on connectivity within large scale brain networks^36^ and their tight association with cortical atrophy^37^. In PWP, OH has been shown to be associated with disrupted connectivity in the rostral prefrontal cortex, anterior cingulate cortex, and other parts of the salience network^38^, known to strongly overlap with the central autonomic network^39,40^. Findings reported here may thus reflect the impact of white matter lesion on this circuitry, although this speculative mechanism needs to be clarified in future research. Orthostatic hypotension has also been shown to alter cerebral perfusion and cerebral hemodynamics^41^, which may selectively impact more vulnerable systems, such as the diseased motor systems in PD. Similarly, in dementia with Lewy bodies (a related alpha-synucleinopathy with high prevalence of OH and dementia which features visuo-spatial deficits), orthostatic hypotension is associated with hypoperfusion of occipito-parietal regions^42^ as well as worse visuo-spatial performance^43^, which are at-risk regions in dementia with Lewy bodies.

Consistent with previous results, our analyses demonstrated that the axial and bradykinesia components of the MDS–UPDRS Part III exam were associated with WMH volume. The association between bradykinesia scores, in particular, and WMH volume was most strongly impacted by OH. Previous literature on the relationship between total WMH burden and the specific sub-sections of the MDS–UPDRS Part III exam have been overall inconsistent. One study found that in de novo PD, an increased WMH burden directly affects the total axial motor score^44^. Another study found an association between deep WMH burden and bradykinesia score, as well as an association of periventricular WMH burden with total bradykinesia and axial motor scores^14^. Multiple studies have similarly found associations between orthostatic hypotension and the Postural Instability Gait Disturbance (PIGD) subtype of Parkinson’s disease, which features higher scores in bradykinesia and axial symptoms and less tremor burden^45^. Our results join these finding by offering additional insight as mediators that may impact the association between WMH volume and specific parkinsonian motor domains.

Several limitations of the current study should be noted. The study’s focus on de novo PD minimized the potentially confounding influence of Levodopa medication regiment on findings, but we acknowledge that this narrowed the ranges of disease durations we could consider. The cross-sectional design used here precluded any insights into how OH impacts progressive changes in the association between WMH volume and motor symptom burden. Future longitudinal studies are needed to determine if the role of OH as a mediating variable between WMH burden and motor dysfunction continues during disease progression. Since very few controls qualified as having OH, we were not able to meaningfully investigate the effect of PD itself on the associations between OH and WMH. This should be addressed in future research with larger control sample sizes.

In conclusion, we found that the association between WMH volume and the burden of motor symptoms in PD, particularly bradykinesia symptoms, is impacted significantly by the presence of OH. These findings may help explain the mechanism underlying the poorer prognosis observed among PWP with OH.

## Supporting information

D'Amico_2026_SM_Final

## Data Availability

All data produced are available online at: https://www.ppmi-info.org/

