## Supplementary material for "Orthostatic hypotension in Parkinson’s disease impacts the association between white matter lesion volume and motor symptom burden": D'Amico_2026_SM_Final

Supplementary Figure 1: *Association between MDS–UPDRS Part III bradykinesia sub-score and WMH burden in PWP with and without OH.*

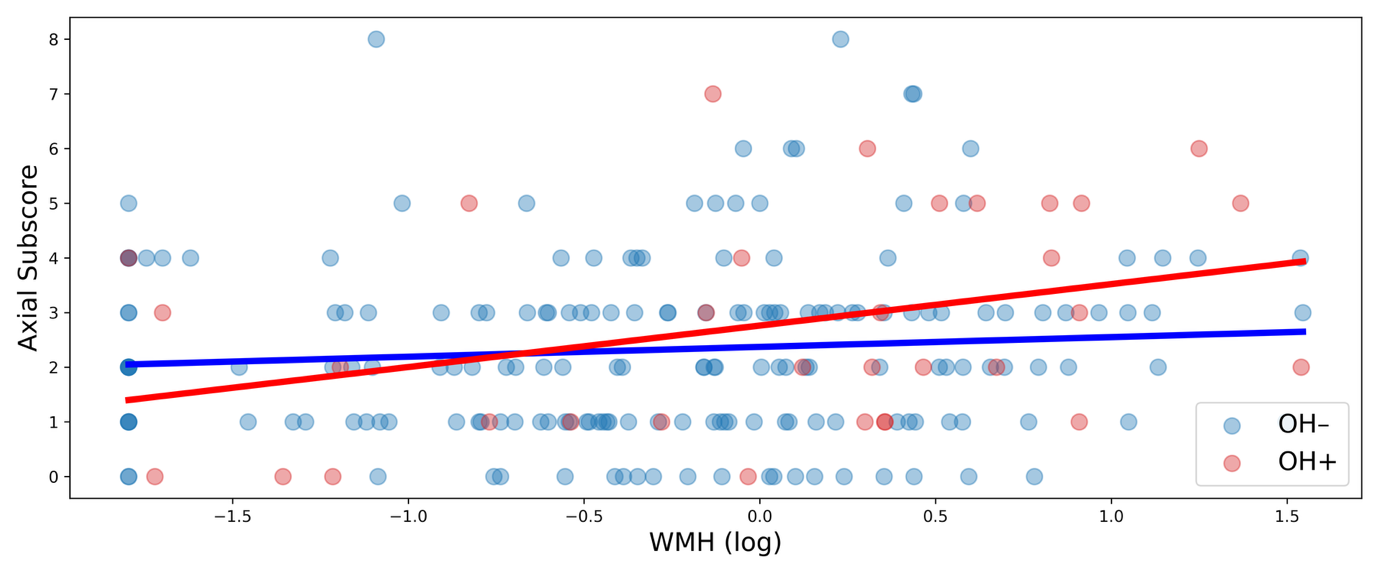

**Supplementary Figure 1: Impact of orthostatic hypotension on associations between white matter hyperintensity volume and axial symptom burden.** An ANCOVA that adjusted for age, sex, and eTIV revealed there are significant differences in the association between the total WMH volume (log-transformed values) and the axial symptom burden (MDS–UPDRS Part III axial sub-scores) in PWP with OH (OH+) compared to those without OH (OH–).

Supplementary Figure 2: *Association between MDS–UPDRS Part III axial sub-score and WMH burden in PWP with and without OH.*

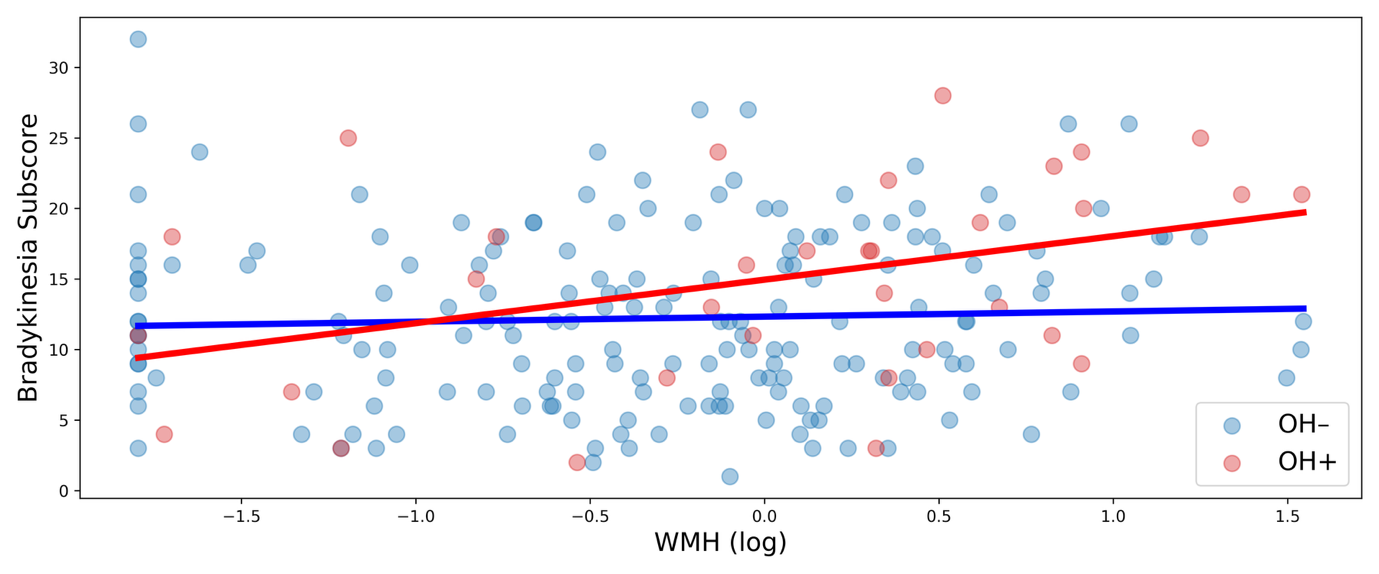

**Supplementary Figure 2: Impact of orthostatic hypotension on associations between white matter hyperintensity volume and bradykinesia symptom burden.** An ANCOVA that adjusted for age, sex, and eTIV of PWP revealed there are significant differences in the association between total WMH volume (log-transformed values) and the bradykinesia symptom burden (MDS–UPDRS Part III bradykinesia sub-scores) in PWP with OH (OH+) compared to those without OH (OH–).

Supplementary Table 1: *Comparison of co-morbidities in PWP with and without OH*

| Condition Classification | Value | N (*OH+/OH–*) | *p* | |
| --- | --- | --- | --- | --- |
| Dermatological | 7.222E-4 | 46 (6/40) | | 0.979 |
| Ophthalmological | 0.326 | 70 (9/61) | | 0.568 |
| ENT | 0.058 | 53 (6/47) | | 0.809 |
| Pulmonary | 0.140 | 35 (4/31) | | 0.708 |
| Cardiovascular | 2.429 | 116 (19/97) | | 0.119 |
| Gastrointestinal | 4.082E-4 | 87 (11/76) | | 0.984 |
| Hepatobiliary | 0.090 | 19 (3/16) | | 0.764 |
| Renal | 2.440 | 18 (5/13) | | 0.118 |
| Gynecological/Urologic | 1.040 | 73 (11/62) | | 0.308 |
| Musculoskeletal | 0.429 | 95 (13/82) | | 0.512 |
| Metabolic/Endocrine | 0.752 | 48 (9/39) | | 0.386 |
| Hemato/Lymphatic | 0.296 | 17 (3/14) | | 0.586 |
| Neurologic | 0.345 | 75 (9/66) | | 0.557 |
| Psychiatric | 0.070 | 59 (9/50) | | 0.792 |
| Allergy/Immunologic | 0.008 | 55 (7/48) | | 0.930 |
| Other | 2.378 | 59 (11/48) | | 0.123 |
| COVID-19 | 0.296 | 21 (3/18) | | 0.586 |

ENT = otolaryngology; Hemato/Lymphatic = Hematological and Lymphatic

Supplementary Table 2: *Pearson’s Correlations among variables of interest in PWP*

| Variable |  | Age | WMH (log) | eTIV (mm^3^) | MDS |
| --- | --- | --- | --- | --- | --- |
| Age | Pearson’s r | –– |  |  |  |
|  | p-value | –– |  |  |  |
| WMH (log) | Pearson’s r | 0.516 | –– |  |  |
|  | p-value | < .001 | –– |  |  |
| eTIV (mm^3^) | Pearson’s r | -0.091 | 0.149 | –– |  |
|  | p-value | 0.183 | 0.028 | –– |  |
| MDS | Pearson’s r | 0.218 | 0.145 | 0.049 | –– |
|  | p-value | 0.001 | 0.032 | 0.471 | –– |

MDS = Total MDS–UPDRS Part III score

Supplementary Table 3: *Pearson’s Correlations among variables of interest in OH+ PWP*

| Variable |  | Age | WMH (log) | eTIV (mm^3^) | MDS |
| --- | --- | --- | --- | --- | --- |
| Age | Pearson’s r | –– |  |  |  |
|  | p-value | –– |  |  |  |
| WMH (log) | Pearson’s r | 0.424 | –– |  |  |
|  | p-value | 0.014 | –– |  |  |
| eTIV (mm^3^) | Pearson’s r | -5.465E-4 | 0.403 | –– |  |
|  | p-value | 0.998 | 0.020 | –– |  |
| MDS | Pearson’s r | 0.270 | 0.386 | 0.095 | –– |
|  | p-value | 0.129 | 0.027 | 0.600 | –– |

MDS = Total MDS–UPDRS Part III score

Supplementary Table 4: *Pearson’s Correlations among variables of interest in OH– PWP*

| Variable |  | Age | WMH (log) | eTIV (mm^3^) | MDS |
| --- | --- | --- | --- | --- | --- |
| Age | Pearson’s r | –– |  |  |  |
|  | p-value | –– |  |  |  |
| WMH (log) | Pearson’s r | 0.519 | –– |  |  |
|  | p-value | < .001 | –– |  |  |
| eTIV (mm^3^) | Pearson’s r | -0.114 | 0.084 | –– |  |
|  | p-value | 0.123 | 0.254 | –– |  |
| MDS | Pearson’s r | 0.179 | 0.061 | 0.035 | –– |
|  | p-value | 0.015 | 0.414 | 0.638 | –– |

MDS = Total MDS–UPDRS Part III score

Supplementary Table 5: *ANCOVA testing the effect of the interaction between OH and WMH volume on total MDS–UPDRS Part III score*

| Cases | Sum of Squares | df | Mean Square | F | p |
| --- | --- | --- | --- | --- | --- |
| Age | 598.954 | 1 | 598.954 | 6.283 | 0.013 |
| Sex | 21.716 | 1 | 21.716 | 0.228 | 0.634 |
| WMH (log) | 200.414 | 1 | 200.414 | 2.102 | 0.149 |
| eTIV (mm^3^) | 57.968 | 1 | 57.968 | 0.608 | 0.436 |
| OH | 354.360 | 1 | 354.360 | 3.717 | 0.055 |
| OH × WMH (log) | 474.424 | 1 | 474.424 | 4.977 | 0.027 |
| Residuals | 20018.117 | 210 | 20018.117 |  |  |

OH Qual = Whether patients qualified as having orthostatic hypotension or not

Supplementary Table 6: *ANCOVA testing the effect of the interaction between OH and WMH volume on total MDS–UPDRS Part III score, adjusting for disease duration*

| Cases | Sum of Squares | df | Mean Square | F | p |
| --- | --- | --- | --- | --- | --- |
| Age | 650.333 | 1 | 650.333 | 6.819 | 0.010 |
| Sex | 40.097 | 1 | 40.097 | 0.420 | 0.517 |
| WMH (log) | 186.019 | 1 | 186.019 | 1.950 | 0.164 |
| eTIV (mm^3^) | 94.158 | 1 | 94.158 | 0.987 | 0.322 |
| OH | 408.728 | 1 | 408.728 | 4.286 | 0.040 |
| OH × WMH (log) | 456.581 | 1 | 456.581 | 4.787 | 0.030 |
| PDDur (mo.) | 248.254 | 1 | 248.254 | 2.603 | 0.108 |
| Residuals | 19741.712 | 207 | 95.371 |  |  |

PDDur = Number of months passed since the participant was diagnosed with PD

Supplementary Table 7: *ANCOVA testing the effect of the interaction between OH and WMH volume on total MDS–UPDRS Part III score, adjusting for symptom duration*

| Cases | Sum of Squares | df | Mean Square | F | p |
| --- | --- | --- | --- | --- | --- |
| Age | 617.834 | 1 | 617.834 | 6.463 | 0.012 |
| Sex | 17.508 | 1 | 17.508 | 0.183 | 0.669 |
| WMH (log) | 201.515 | 1 | 201.515 | 2.108 | 0.148 |
| eTIV (mm^3^) | 57.816 | 1 | 57.816 | 0.605 | 0.438 |
| OH | 396.434 | 1 | 396.434 | 4.147 | 0.043 |
| OH × WMH (log) | 466.050 | 1 | 466.050 | 4.876 | 0.028 |
| SymptDur (mo.) | 202.831 | 1 | 202.831 | 2.122 | 0.147 |
| Residuals | 19787.136 | 207 | 95.590 |  |  |

SymptDur = Duration of Parkinsonian symptoms

Supplementary Table 8: *ANCOVA testing the effect of the interaction between OH and WMH volume on total MDS–UPDRS Part III score, adjusting average DaT*

| Cases | Sum of Squares | df | Mean Square | F | p |
| --- | --- | --- | --- | --- | --- |
| Age | 499.459 | 1 | 499.459 | 5.388 | 0.021 |
| Sex | 40.803 | 1 | 40.803 | 0.440 | 0.508 |
| WMH (log) | 192.407 | 1 | 192.407 | 2.075 | 0.151 |
| eTIV (mm^3^) | 56.183 | 1 | 56.183 | 0.606 | 0.437 |
| OH | 323.326 | 1 | 323.326 | 3.488 | 0.063 |
| OH × WMH (log) | 553.515 | 1 | 553.515 | 5.971 | 0.015 |
| DaT Average | 642.880 | 1 | 642.880 | 6.935 | 0.009 |
| Residuals | 19375.237 | 207 | 92.704 |  |  |

Supplementary Table 9: *ANCOVA testing the effect of the interaction between OH and WMH volume on MDS–UPDRS Part III bradykinesia sub-score*

| Cases | Sum of Squares | df | Mean Square | F | p |
| --- | --- | --- | --- | --- | --- |
| Age | 170.693 | 1 | 170.693 | 4.554 | 0.034 |
| Sex | 80.984 | 1 | 80.984 | 2.160 | 0.143 |
| WMH (log) | 81.172 | 1 | 81.172 | 2.165 | 0.143 |
| eTIV (mm^3^) | 37.009 | 1 | 37.009 | 0.987 | 0.322 |
| OH Qual | 155.977 | 1 | 155.977 | 4.161 | 0.043 |
| OH × WMH (log) | 192.710 | 1 | 192.710 | 5.141 | 0.024 |
| Residuals | 7872.007 | 210 | 37.486 |  |  |

Supplementary Table 10: *ANCOVA testing the effect of the interaction between OH and WMH volume on MDS–UPDRS Part III axial sub-score*

| Cases | Sum of Squares | df | Mean Square | F | p |
| --- | --- | --- | --- | --- | --- |
| Age | 43.738 | 1 | 43.738 | 15.657 | < .001 |
| Sex | 3.607 | 1 | 3.607 | 1.291 | 0.257 |
| WMH (log) | 4.105 | 1 | 4.105 | 1.469 | 0.227 |
| eTIV (mm^3^) | 1.363 | 1 | 1.363 | 0.488 | 0.486 |
| OH | 0.352 | 1 | 0.352 | 0.126 | 0.723 |
| OH × WMH (log) | 8.636 | 1 | 8.636 | 3.091 | 0.080 |
| Residuals | 586.626 | 210 | 2.793 |  |  |
